# Analysis and Mitigation of Equipment-induced Shortcuts in AI Models for Laparoscopic Cholecystectomy

**DOI:** 10.64898/2026.04.22.26351545

**Authors:** Sergey Protserov, Anastasiia Repalo, Pouria Mashouri, Jaryd Hunter, Caterina Masino, Amin Madani, Michael Brudno

## Abstract

Machine learning models have seen a lot of success in medical image segmentation domain. However, one of the challenges that they face are confounders or shortcuts: spurious correlations or biases in the training data that affect the resulting models. One example of such confounders for surgical machine learning is the setup of surgical equipment, including tools and lighting. Using the task of identification of safe and dangerous zones of dissection in laparoscopic cholecystectomy images and videos as a use-case, we inspect two equipment-induced biases: the presence of surgical tools in the field of view and the position of lighting. We propose methods for evaluating the severity of these biases and augmentation-based methods for mitigating them. We show that our tool bias mitigations improve the models’ consistency under tool movements by 9 percentage points in the most inconsistent cases, and by 4 percentage points on average. Our lighting bias mitigations help reduce fraction of true dangerous zone pixels that may be predicted as safe under light changes from 5% to 1.5%, without compromising segmentation quality.

## 1. Introduction

Surgical complications represent a substantial burden on global health-care systems, contributing significantly to patient morbidity, mortality, and economic costs. Research indicates[1, 2] that a significant portion of surgical adverse events is caused by errors in perception and decision-making among surgeons.

The integration of artificial intelligence and machine learning technologies into surgical practice offers a promising approach to addressing these challenges by providing real-time decision support to surgical teams. Advanced deep learning algorithms have demonstrated[3] remarkable capabilities in surgical computer vision tasks, achieving performance comparable to that of expert surgeons for the tasks of anatomical structure identification.

In recent years, deep learning machine models based on convolutional [4] and transformer [5] neural networks have seen much success in the field of medical image segmentation, as evidenced by multiple systematic reviews [6, 7, 8, 9, 10, 11, 12, 13]. However, clinical utility of such models is not necessarily directly reflected by segmentation evaluation metrics, such as Dice score, precision and recall. Deployment of these models to clinical environments and their real-world usefulness are hindered by confounders present in the input images [14], allowing the model to demonstrate high performance on held-out test data that does not generalize to other settings. Some examples of such confounders are information that allows the model to distinguish between different hospital systems or departments [15] which have different prevalence of outcome of interest, or location of the object in the field of view being correlated with the segmentation label [16].

In this paper we consider the problem of identifying safe and dangerous zones of dissection during laparoscopic cholecystectomy (LC). While prior work[17, 18] has shown that computer vision techniques can have broad applicability to this problem, we have identified two medical equipment-related confounders that are present in training and testing datasets, and affect models developed using this data. Specifically, medical tool location in the field of view and the direction of lighting are both highly correlated with safe and dangerous zone locations, due to how this surgery is performed. The tool tips of dissection instruments (scalpels, cauterizers) are most often found in the safe zone, as this is exactly where the surgeons perform dissections.

Similarly, lighting is typically directed to the same zone of interest. Direction of lighting may be further correlated with the tool location, potentially complicating the relationship between tool, lighting and zones of interest.

In this work we propose methods for evaluating the extent to which a given model is affected by these biases, and show that the models trained on biased datasets are indeed confounded by tool and lighting locations. We further propose augmentation-based strategies for mitigation of these issues, based on pasting in images of surgical instruments in random positions and random local lighting adjustments to break the correlations with safe/dangerous zone location. We show that the best tool bias mitigation result is achieved when the tool pasting augmentation is applied twice per input image during training, and this helps improve the models’ consistency under tool movements by 9 percentage points in the most inconsistent cases, and by 4 percentage points on average, and that tool texture plays a significant role in these augmentations. We also found that the worst case for lighting bias is when the lighting is directed to the regions inside the predicted safe zone, and in such cases our mitigations help reduce the fraction of true dangerous zone pixels that may be predicted as safe zone under different lighting conditions from 5% to 1.5%. Altogether, we demonstrate that our mitigation strategies indeed help train unbiased models, without compromising segmentation quality.

## 2. Background work

Studies that consider the computer vision tasks related to laparoscopic surgery include EndoNet [19], where the authors develop a model for surgery phase recognition and tool detection in laparoscopic videos, DeepCVS [20], where the authors develop a model for highlighting hepatocystic anatomy and assessing whether the critical view of safety (CVS) has been established during laparoscopic cholecystectomy, and the study [17], where the authors develop models for highlighting hepatocystic anatomy and safe and dangerous zones of dissection during laparoscopic cholecystectomy. More recently Surgestures [21] focused on identifying the surgeon movements during laparoscopic cholecystectomy that can be used to quantify and summarize operative patterns of surgeons with different levels of expertise, while Protserov et al.[18] further improved upon results from [17] and proceeded to deploy an optimized version of the model for safe/dangerous zone highlighting. Our work builds upon these publications, however focuses specifically on the biases introduced by medical equipment setup in the field of view. Finally, Zheng et al [22] describe a generative model-based system for improving the quality of laparoscopic videos by reducing image blur and removing smoke and camera fog. They demonstrate that the use of this system helps reduce the operation pause time and surgeon anxiety.

Another broad field of research relevant to this study is shortcut learning [14]. Confounders make models learn spurious correlations (shortcuts) in training data, instead of meaningful features. They are particularly insidious as they are likely to be also present in the test set built via a random split of the training data, leading to overconfidence in the performance of the model. One noteworthy example of such shortcuts is described in [15], where the authors found that convolutional neural networks trained for pneumonia screening from chest X-rays fail to generalize to external sites, while being able to predict which hospital system and even which hospital department a given image comes from. Analysis of activation maps showed that when predicting the source of a given image, the model “pays attention” to image corners, and laterality tokens specifically. The main finding of this paper is that when different hospital systems have different levels of pneumonia prevalence, the models can indirectly leverage this information to produce confounded predictions. The work [16] further studies shortcut learning in the context of medical image segmentation on two different datasets, demonstrating that presence of text and calipers in the fetal ultrasound images provides shortcuts for organ segmentation, and that centered location of skin lesions in the skin lesion segmentation dataset provides shortcuts that cause the models to under-perform if the lesion is not located at the center of the image at test time. The study [23] focuses on the shortcut in classifier models for skin cancer prediction which learn that in the ISIC dataset color calibration charts occur only in benign images, but not in malignant ones. The authors demonstrate the existence of this bias in trained models by artificially inserting these charts into malignant images, and inpainting them away in benign images. They further propose a de-biasing strategy based on inpainting the charts with benign skin pixels in biased images. We are not aware of any studies that focus specifically on shortcuts or confounders in surgical videos.

To reduce the impact of shortcuts, general-purpose shortcut learning mitigation strategies are available, such as Deep Feature Reweighting (DFR, [24]), which amounts to re-training a model’s last layer on unbiased data, and MARG-CTRL [25] family of methods, which offers shortcut learning mitigation through various modifications of cross-entropy loss used during training.

## 3. Methods

### 3.1. Ethics statement

This research uses de-identified patient data, and has been approved by the University Health Network (UHN) Research Ethics Board, REB#20-5349.

### 3.2. Data

In this research, we use a dataset previously described in [18] as Dataset 1, that consists of 2627 images from laparoscopic camera extracted from 289 LC videos across 136 different institutions from 37 countries and all continents, with pixel-level annotations of safe and dangerous zones of dissection available for every image. Safe zones correspond to the areas where a surgeon should perform dissections, while dangerous zones are locations where dissection is likely to result in an injury to the patient. The dataset was split into training, validation and testing subsets on a per-video basis, with 220, 43 and 45 videos assigned to the corresponding sets. An example of annotated image from this dataset is shown in Figure 1.

**Figure 1:**
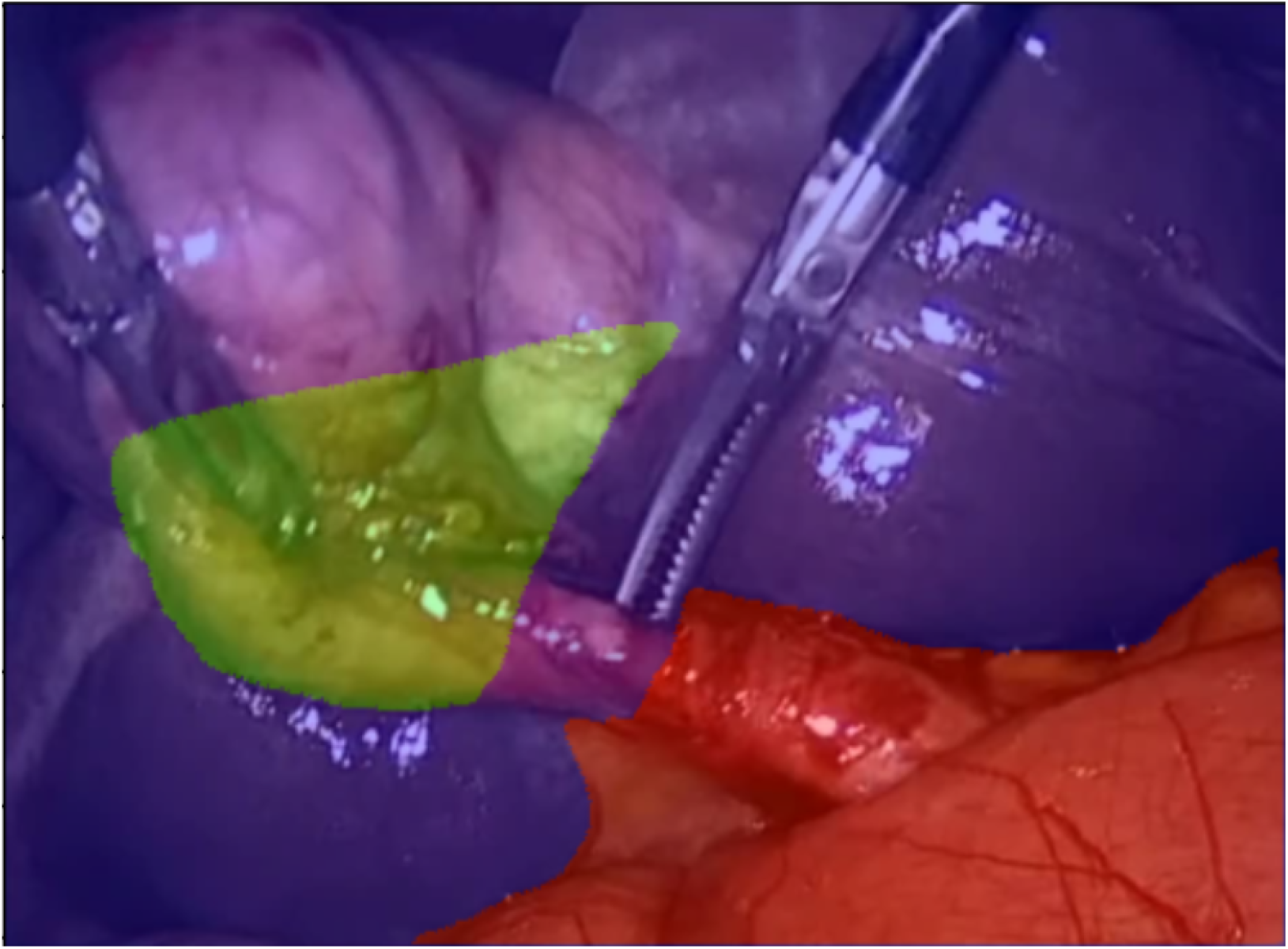
An example annotated image from the LC dataset. Red highlight shows dangerous zone, green – safe zone, blue – background

### 3.3. Tool bias

#### 3.3.1. Problem statement

When conducting LC, the surgeons use a variety of surgical tools and instruments to perform a number of tasks, including dissecting, spreading, cutting or burning through tissues. Examples include scalpels, graspers and cauterization tools. If we consider the problem of identifying and highlighting safe and dangerous zones of dissection, it is clear that there is likely to be strong correlation between the positions of tools and the safe zone locations in any dataset for this problem (experienced surgeons are likely to only use tools in safe zones). Because of that, we hypothesize that models trained on such data will be biased by presence of the tools in the field of view, and may provide erroneous predictions depending on the actual location of the surgical tool. This would have the potential of being harmful if the tool is used by a less experienced surgeon, as the safe zone prediction will potentially “follow” the surgical tools.

#### 3.3.2. Evaluation

Intuitively, a perfectly unbiased model would output the exact same predictions for two otherwise similar input frames that only differ in the positions of surgical tools. Our proposed tool bias evaluation methodology builds upon this idea. We manually identified 23 short video fragments in the testing subset of videos, such that most of the movement in these fragments is attributed to surgical tools, and the background is mostly static. We then used Track Anything [26] program to interactively create frame-by-frame pixel-level annotations of surgical tools in these videos, while manually verifying every annotation. Then, for every such short video, we considered all frames with a step of 100 ms, and all possible pairs of these frames. We used Hausdorff distance between tool locations in the two frames, and mean squared distance between all non-tool pixels to identify frame pairs where the tools are relatively far apart, while the background is largely unchanged. For every video,we selected 50 frame pairs that best satisfied these criteria. To evaluate a model for tool bias, we obtain the model’s predictions on all selected frames, and consider the predictions’ consistency between frames within every selected pair by measuring class average (average over the three segmentation classes: safe, dangerous and background) of Dice score between predictions obtained for both frames. We chose to use Dice score for this purpose because it is symmetric with respect to which image is considered as ground truth vs which is a predicted mask; this is important since the two frames within a pair are interchangeable. Our goal is to validate whether model’s predictions change due to the movement of the tool; the more they change, the more a model is affected by the presence of surgical instruments. Note that ground truth is not used for tool bias assessment because we don’t have ground truth for most frames in the videos, hence we use consistency between similar frames as a proxy.

#### 3.3.3. Mitigations

To mitigate the tool bias, we used 215 images of surgical tools extracted from the training subset of videos, and defined a tool pasting augmentation, which pastes tools in random positions in the image during training, properly adjusting their size to the size of the target image, and with random rotations. We hope that pasting surgical tools in random places in the input image will break the correlation between tool position and safe zone location, leading to models that are more robust in the presence of surgical instruments. Some examples of images augmented in this manner are presented in Figures 2a-c.

**Figure 2:**
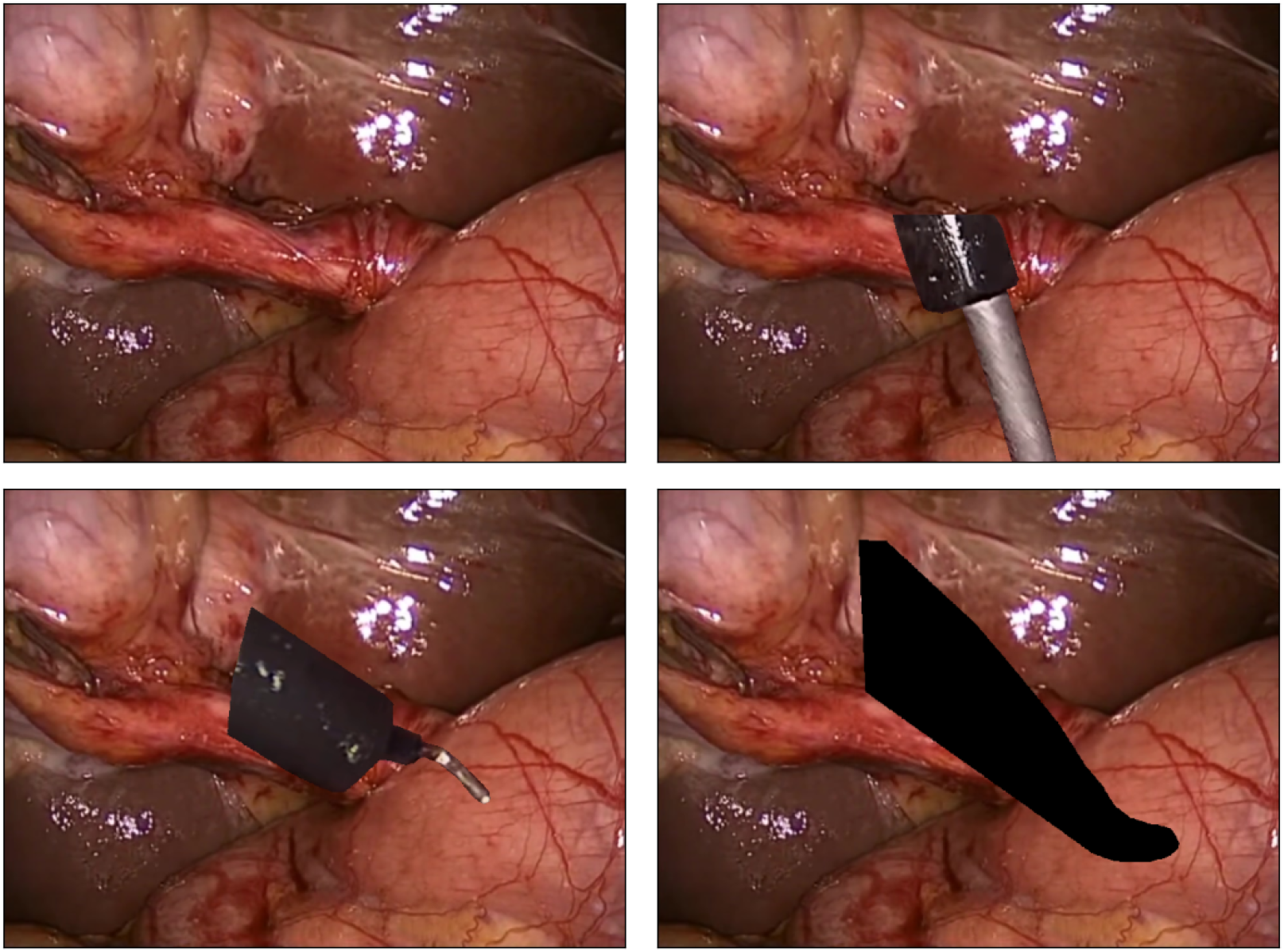
Examples of images augmented with our tool pasting augmentation. a.) Original image, without tool augmentation, b.) Augmented image 1, c.) Augmented image 2, d.) Augmented image 3; Here pasted tool is filled with black color to validate whether the texture of the tool is significant

To simulate presence of multiple tools in the field of view and make our augmentation stronger, we also tried applying the tool pasting augmentation several times to the same image, with 1 to 3 tools being pasted into every image, depending on a model training hyperparameter.

It is important to note that tool pasting augmentation also has an implicit effect of covering large sections of the original input, which may serve as a strong regularization technique in itself. To study the effect obscuring parts of the image may have on the model, we also consider the augmentation mode where the pasted tool is filled completely with black color (Figure 2d). On average, applying tool pasting augmentation once obstructs 6% of the input image, and applying it twice obstructs 11.5% of the image.

### 3.4. Lighting bias

#### 3.4.1. Problem statement

During LC, the light mounted on the laparoscopic camera is typically directed to the region of interest, where the dissection is being performed. Therefore, there is likely to be a correlation between direction of lighting and safe zone locations. This bias is also likely to be learned by the model, and may impact the performance of a system in a real-world deployment, especially when used by less experienced surgeons.

#### 3.4.2. Evaluation

To evaluate to what extent a given model suffers from lighting bias with a given input image, we first obtain the model’s prediction on this image, and then simulate additional lighting in different parts of the image. To study the effect directing lighting to different locations might have on predictions, we simulate lighting centered at different points in the predicted safe zone only, in the predicted dangerous zone only, or anywhere in the image. For every input image, for every lighting simulation mode, we created 50 transformed images. We then assess by how much predictions on the images with simulated lighting differ from the prediction on the original image. The intuition is that a model that is robust to lighting changes should have high consistency. We conduct this experiment on all images in the test subset, and aggregate the results across the images.

#### 3.4.3. Mitigations

Similarly to the idea of our tool pasting augmentation described in Section 3.3.3, we implement a lighting augmentation that locally increases brightness of a random region within the image. The input image is converted from RGB to LAB color space, and a Gaussian blob is added to the L-channel with a center at a random position. The image is then converted back to RGB. For the purposes of evaluation, the intensity of the brightest point (center) of the Gaussian lighting blob is selected randomly between 20 and 50 L-channel points (which range from 0 to 100), for the purposes of augmentation it is fixed at 50 L-channel points. Some examples of images augmented in this manner are presented in Figure 3.

**Figure 3:**
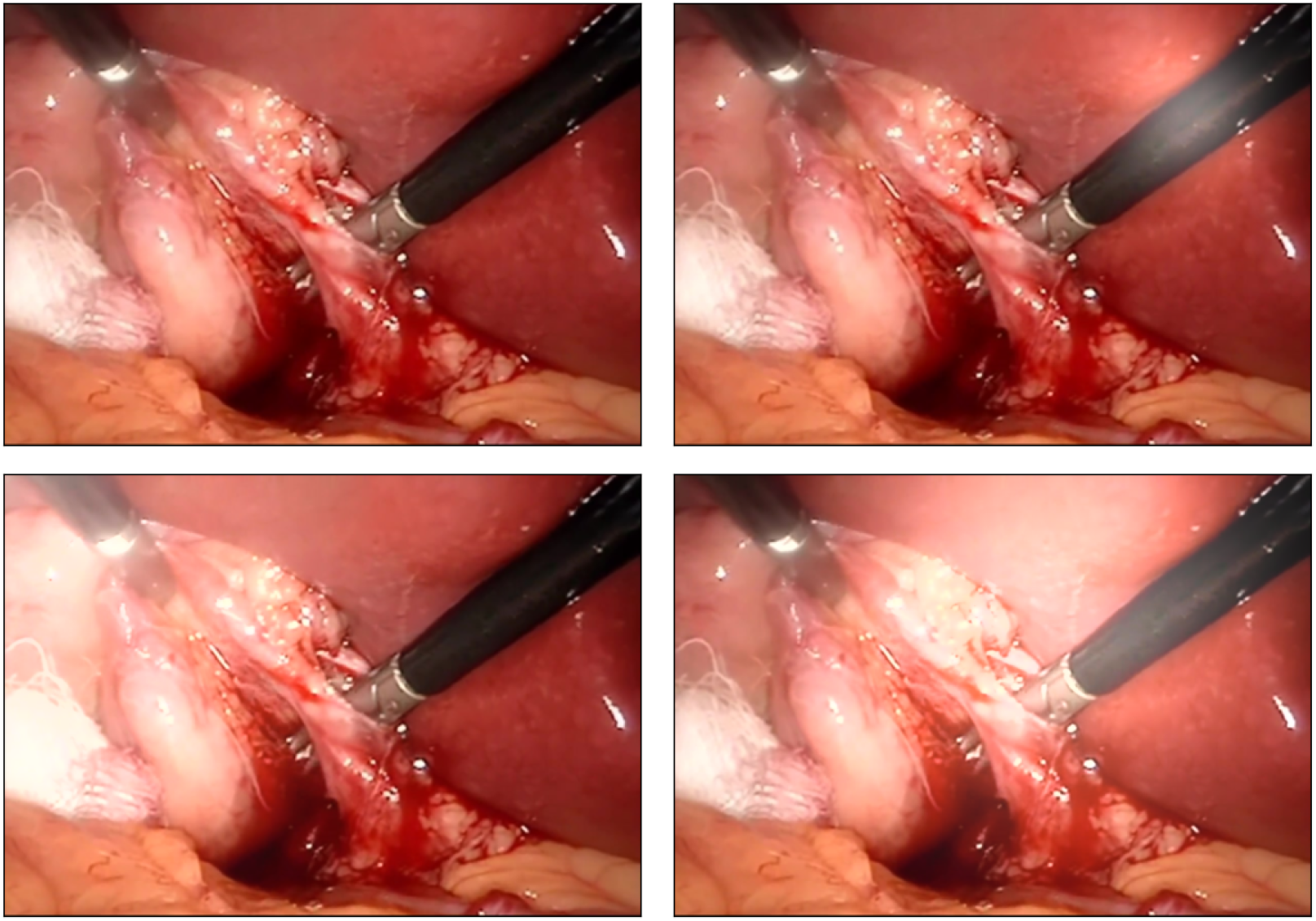
Examples of images augmented with our lighting augmentation. a.) Original image, without lighting augmentation, b.) Augmented image 1, c.) Augmented image 2, d.) Augmented image 3

## 4. Results

### 4.1. Baseline setup

We used SegFormer [27] architecture, and followed the standard machine learning practice of training multiple models with different hyperparameter sets to identify the best model with respect to value of the utilized loss function, cross-entropy, on the validation set. The best model selected in this manner is then used for evaluation on the test set. In our earlier work [18] we showed that SegFormer performs on par with U-Net[28], another well-established semantic segmentation architecture. We resize every image to a height of 128 pixels while preserving the aspect ratio. Our previous work with this dataset showed that this resolution is sufficient. The brightest 100×100 square within the image is selected, and the entire image is standardized so that this square has zero per-channel mean and unit per-channel standard deviation. This strategy is used instead of “regular” standardization, because some inputs have black borders (see Figure 4a) which would distort computations of mean and standard deviation if they were performed over the entire image.

**Figure 4:**
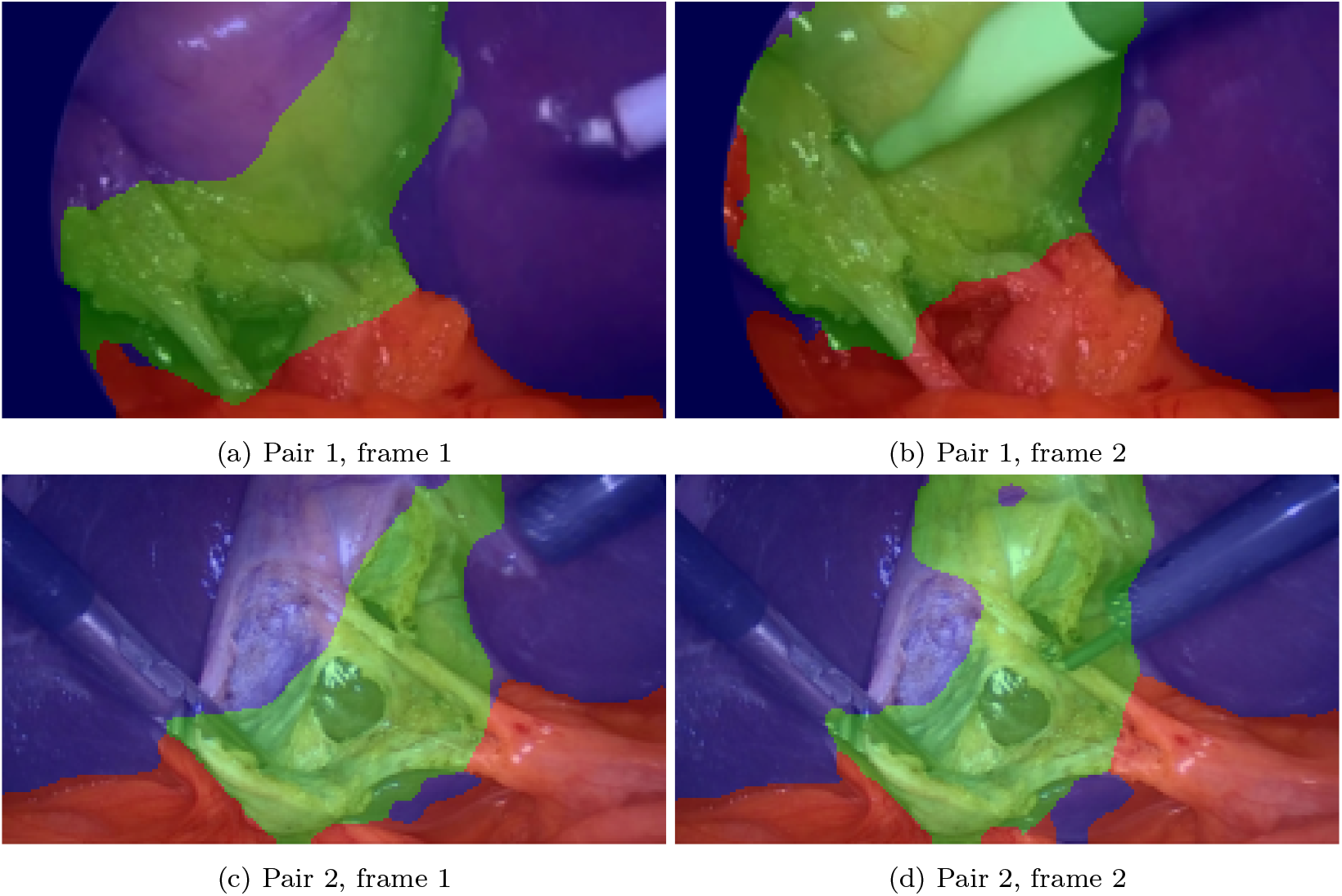
Prediction inconsistencies on two pairs of frames, where frames within each pair are similar except for tool positions. Red highlights predicted dangerous zone, green – predicted safe zone, blue – predicted background. In both frames the presence of the surgical instrument has resulted in a larger predicted “safe” zone. Ground truth segmentations are not available for these frames.

### 4.2. Baseline tool bias

For our best model trained without tool pasting or lighting augmentations, the per-video consistency scores (described in Section 3.3.2), averaged over all selected frame pairs within a video, range between 0.67 and 0.97, with a mean of 0.87 and standard deviation of 0.075, which shows that the model is sensitive to surgical tool position within the field of view. Examples of the prediction inconsistency on two pairs of frames, where frames within a pair are similar except for tool position, are presented in Figure 4.

### 4.3. Tool bias mitigations

Our findings aggregated over videos are presented in Figure 5. They show that for 18 out of 23 videos the model trained with one iteration of tool augmentations is more consistent than the baseline model, and that for 19 out of 23 videos the model trained with two iterations of tool augmentations is more consistent than the baseline model. Also, for 17 out of 23 videos two-iteration model is more consistent than one-iteration model.

**Figure 5:**
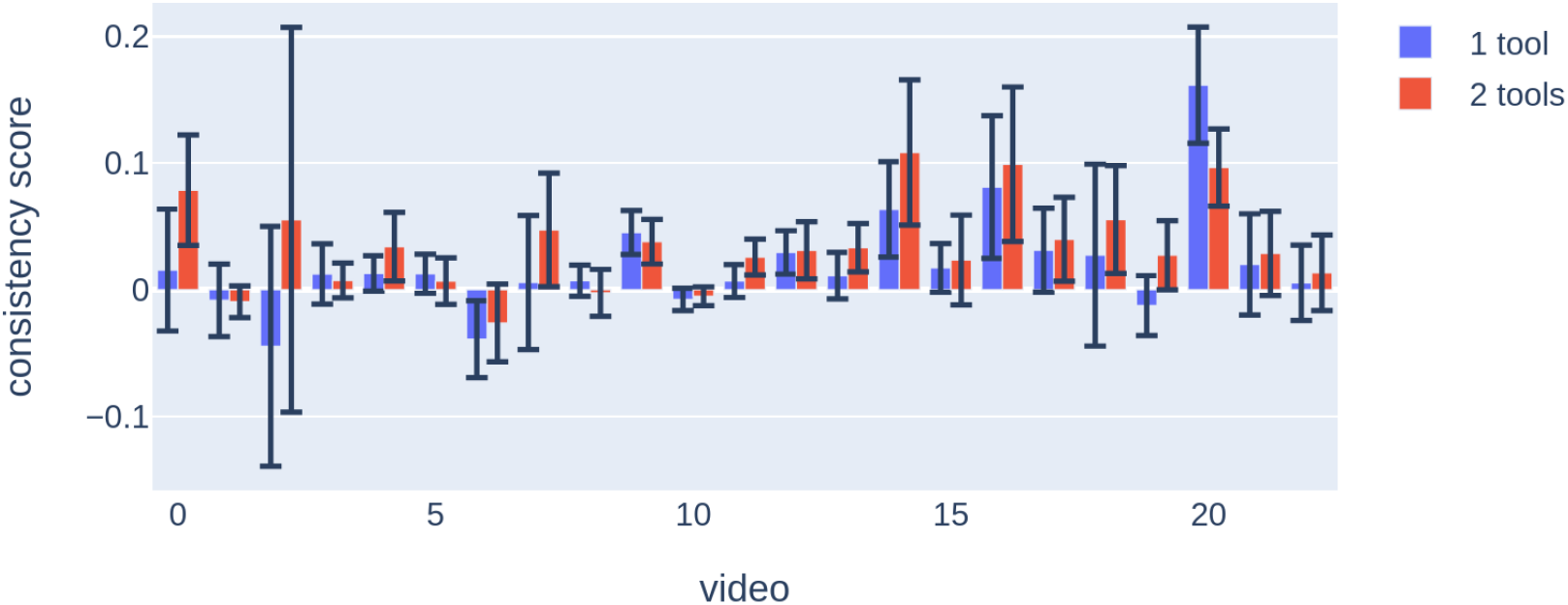
Per-video difference in consistency score of the models trained with one (blue) or two (red) iterations of tool pasting augmentation versus the baseline model, for every short video fragment. Error bars show standard deviation over 50 frame pairs

To validate our claims statistically, we conducted three one-sided t-tests with paired samples, with the null hypotheses being “average per-video consistency scores for the model without any mitigations are not less than scores for the model with one iteration of tool pasting augmentation”, “average per-video consistency scores for the model without any mitigations are not less than scores for the model with two iterations of tool pasting augmentation”,”average per-video consistency scores for the model with one iteration of tool pasting augmentation are not less than scores for the model with two iterations of tool pasting augmentation”, with n=23 for every test. The resulting p-values are, respectively, 0.016, 4.65 · 10^−5^ and 0.014, indicating significance and rejection of the null hypothesis in all three cases.

Our findings (summarized over all 23 videos) are presented in Table 1.

**Table 1:** A summary of consistency scores over all videos, for different tool bias mitigation strategies. Applying tool pasting augmentation once or twice improves the model’s robustness, while merely covering parts of the image with tool-shaped black objects does not result in any improvement. No additional benefit is achieved from pasting three tools

| mitigation strategy | min | max | mean ± std |
| --- | --- | --- | --- |
| no augmenations | 0.668 | 0.970 | $0.873 \pm 0.075$ |
| paste 1 black shape | 0.674 | 0.966 | $0.876 \pm 0.073$ |
| paste 1 real tool image | 0.698 | 0.971 | $0.893 \pm 0.062$ |
| paste 2 black shapes | 0.652 | 0.973 | $0.879 \pm 0.084$ |
| paste 2 real tool images | 0.765 | 0.965 | $0.908 \pm 0.052$ |
| paste 3 real tools images | 0.780 | 0.977 | $0.897 \pm 0.048$ |
| DFR | 0.667 | 0.945 | $0.872 \pm 0.073$ |
| MARG-CTRL | 0.714 | 0.961 | $0.897 \pm 0.053$ |

For our best model trained with a tool pasting augmentation applied once for every input image during training, the per-video consistency scores range between 0.7 (was 0.67 for the baseline model) and 0.97 (was 0.97), with a mean of 0.89 (was 0.87) and standard deviation of 0.062 (was 0.075). For our best model trained with a tool pasting augmentation applied twice for every input image, the per-video consistency scores further improved to range between 0.76 and 0.96, with a mean of 0.91 and standard deviation of 0.052. This demonstrates that applying tool pasting augmentation helps improve prediction consistency scores, and that applying the augmentation twice for every image amplifies the effect. For every selected short video fragment, for every selected pair of frames, we compared consistency of models trained with one and two iterations of tool pasting augmentation against the baseline model by subtracting the per-frame-pair consistency scores and averaging over the frame pairs in every video.

To study the effect increasing the amount of iterations of tool pasting augmentation has on tool bias, and to distinguish between the de-biasing effect from tool pasting and the regularization effect from obstructing parts of the image, we also conducted experiments with three iterations of tool pasting, as well as one or two iterations of pasting tools filled with black color. They show that making more than two iterations of tool pasting augmentation does not provide any further benefit, and that tool texture actually plays a significant role, as merely obstructing parts of the input image with tool-shaped black objects does not help reduce tool bias at all.

We also implemented two pre-existing shortcut learning mitigation methods: DFR [24] and MARG-CTRL [25] (marg-log and SD variants), with proper hyperparameter optimization. DFR did not provide any improvement in consistency, while MARG-CTRL still lagged behind the “two tools” version of our augmentation (Table 1). Another advantage of our approach is that MARG-CTRL’s derivation relies on the model being trained with cross entropy loss and is not necessarily compatible with, e.g. the Dice loss that is frequently used in segmentation settings, and also with object detection problem formulation. Our augmentation is data-driven and loss- and task-agnostic.

To statistically compare our method against MARG-CTRL, we took our strongest augmentation (“two tools” version), and fit a linear mixed-effects model to consistency scores calculated between pairs of video frames. For these purposes, we used lme4 library [29]. The mixed effect model included a fixed effect for the method of correction, and random effects for both the video and frame pair. Our method demonstrated significantly higher consistency scores than MARG-CTRL (p-value=0.0148).

In Figure 6 we show the predictions of the model trained with two iterations of tool pasting augmentations on the same pairs of frames as in Figure It can be seen that this model is much more consistent than the baseline model.

**Figure 6:**
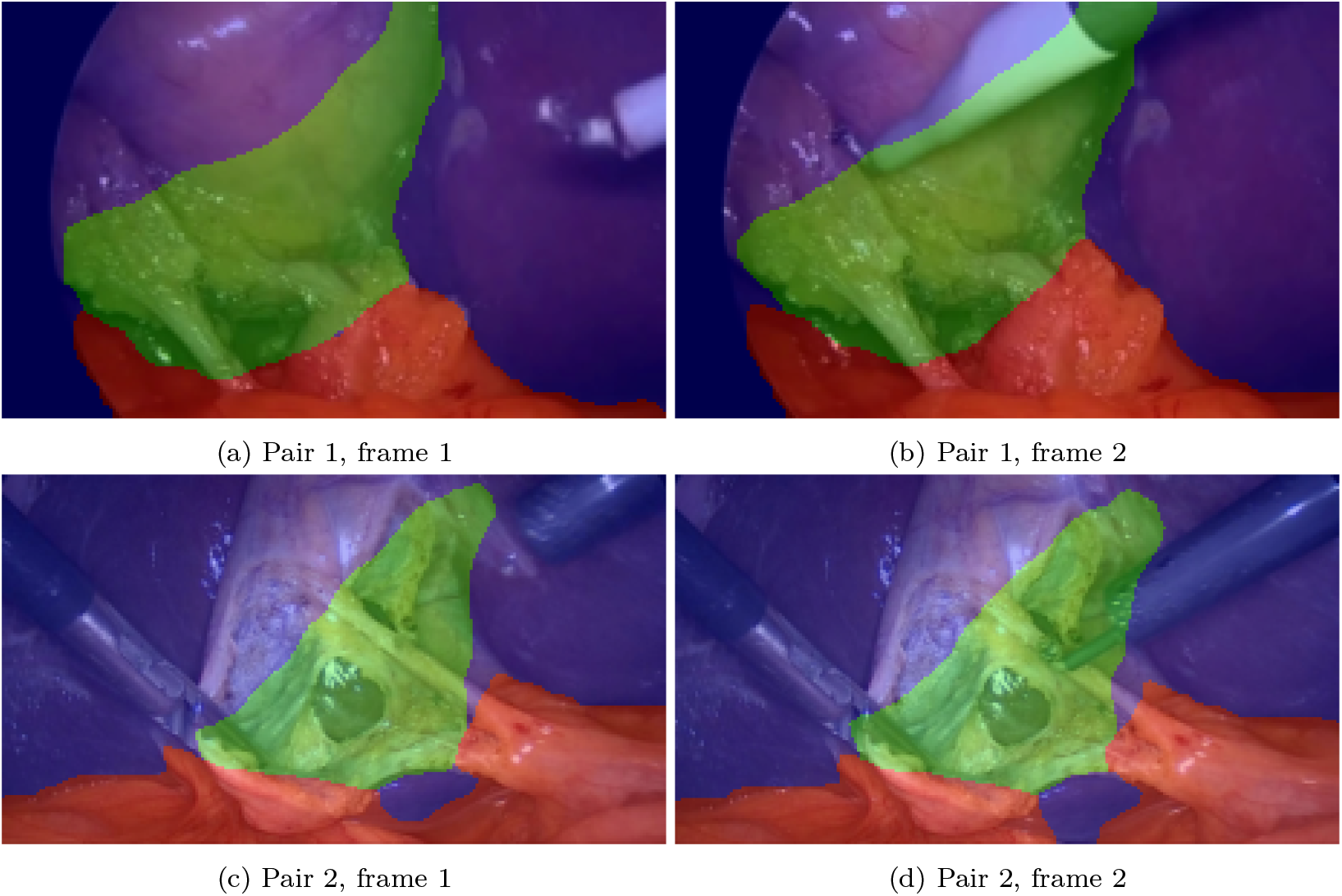
Predictions of the model trained with two iterations of tool augmentation on the same pairs of frames as were shown in Figure 4. Red highlights predicted dangerous zone, green – predicted safe zone, blue – predicted background. This model shows much better consistency than the baseline model (Figure 4). Ground truth segmentations are not available for these frames.

### 4.4. Validation at scale

While the results presented in the previous section clearly demonstrate that our proposed mitigation methods help improve model consistency under tool movements, obtaining these results required manual data selection and annotation. To further validate the improvements of our augmented model compared to the baseline, as well as demonstrate the ability of our evaluation methodology to scale beyond manually curated data, we acquired two extra videos from CholecTrack20 dataset[30], and performed the following steps:

1. Trained a model for semantic segmentation of surgical tools on the Endoscapes[31] dataset
2. Used this model to obtain tool segmentations for the above-mentioned video segments
3. Using a sliding window of 5 seconds over the relevant parts of these videos, considered pairs of frames within every window
4. Computed Hausdorff distances between tools, MSEs between non-tool pixels, and safe/dangerous zone segmentation consistencies of the baseline and augmented models for the frame pairs
5. Considered the frame pairs from the two videos together, binned them by MSE and plotted consistency improvement of the augmented model versus baseline depending on Hausdorff distances between tools in the frame pairs

The resulting plot (Figure 7) shows by how many percentage points our augmented model is more consistent than the baseline, as the tool movement between frames in a pair grows. This plot shows that the improvement in consistency is always positive and that it grows as the Hausdorff distance between tool positions grows. This is in agreement with our theory that larger discrepancies in tool locations should make it more challenging for the model to stay consistent, and that our mitigation strategies help ensure consistency.

**Figure 7:**
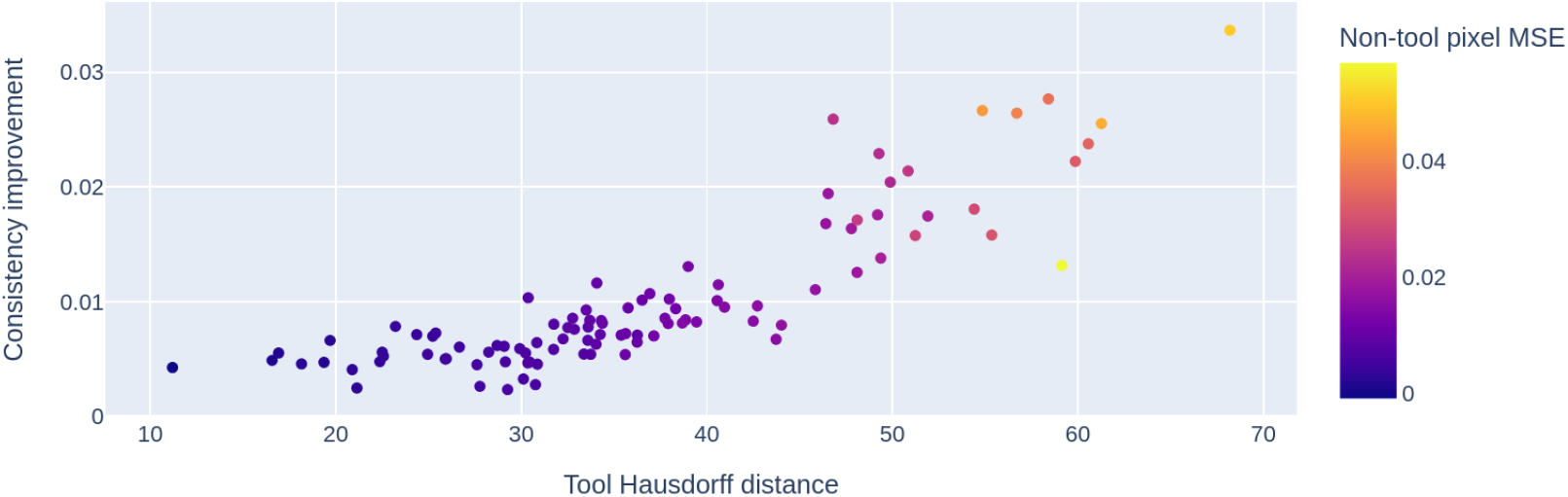
Safe/dangerous zone segmentation consistency improvement of tool-augmented model versus baseline, binned over non-tool pixel MSE and plotted against tool position Hausdorff distance

One could argue that numerical improvements of consistency may be misleading, because the model may be very consistent, but consistently wrong. In Section 4.7 we show that this is not the case for our augmented models, as they perform roughly on the same level as baseline models in terms of segmentation metrics.

Another takeaway from these experiments is that the surgical tool semantic segmentation model trained on Endoscapes dataset can be further used to extract more tool images and remove the need for manual tool annotation prior to using them for augmentation.

### 4.5. Baseline lighting bias

As described in Section 3.4.2, to evaluate lighting bias we assess how much the model’s predictions change when we consider predictions on the original input image to be “ground truth”, and compare predictions on images with local lighting adjustments against them. We found that the most drastic changes occur when the center of the simulated lighting blob is in the predicted safe zone, and further consider this case specifically. Figure 8a shows row-normalized confusion matrix (computed for every original image with all its transformations, then averaged over all original images), where every entry in position *c*_*i*_, *c*_*j*_ shows what proportion of pixels that originally belonged to class *c*_*i*_ belong to class *c*_*j*_ in the transformed image. These results show that on average, 5% of the pixels originally predicted as a dangerous zone are classified as safe due to lighting adjustment. This is a dangerous mistake to make. Further, lighting adjustment causes another 5% of pixels originally belonging to dangerous zone to be predicted as background. Figure 8b shows fully-normalized confusion matrix (computed for every original image with all its transformations, then averaged over all original images), where every entry in position *c*_*i*_, *c*_*j*_ shows what proportion of pixels originally belonged to class *c*_*i*_ and belong to class *c*_*j*_ in the transformed image. Fully-normalized confusion matrices are used to highlight zone size changes. Baseline model that does not use augmentation shows higher changes in zone sizes: dangerous zone shrinks by 7%, while safe zone and background grow by 2% and 1.5% respectively.

**Figure 8:**
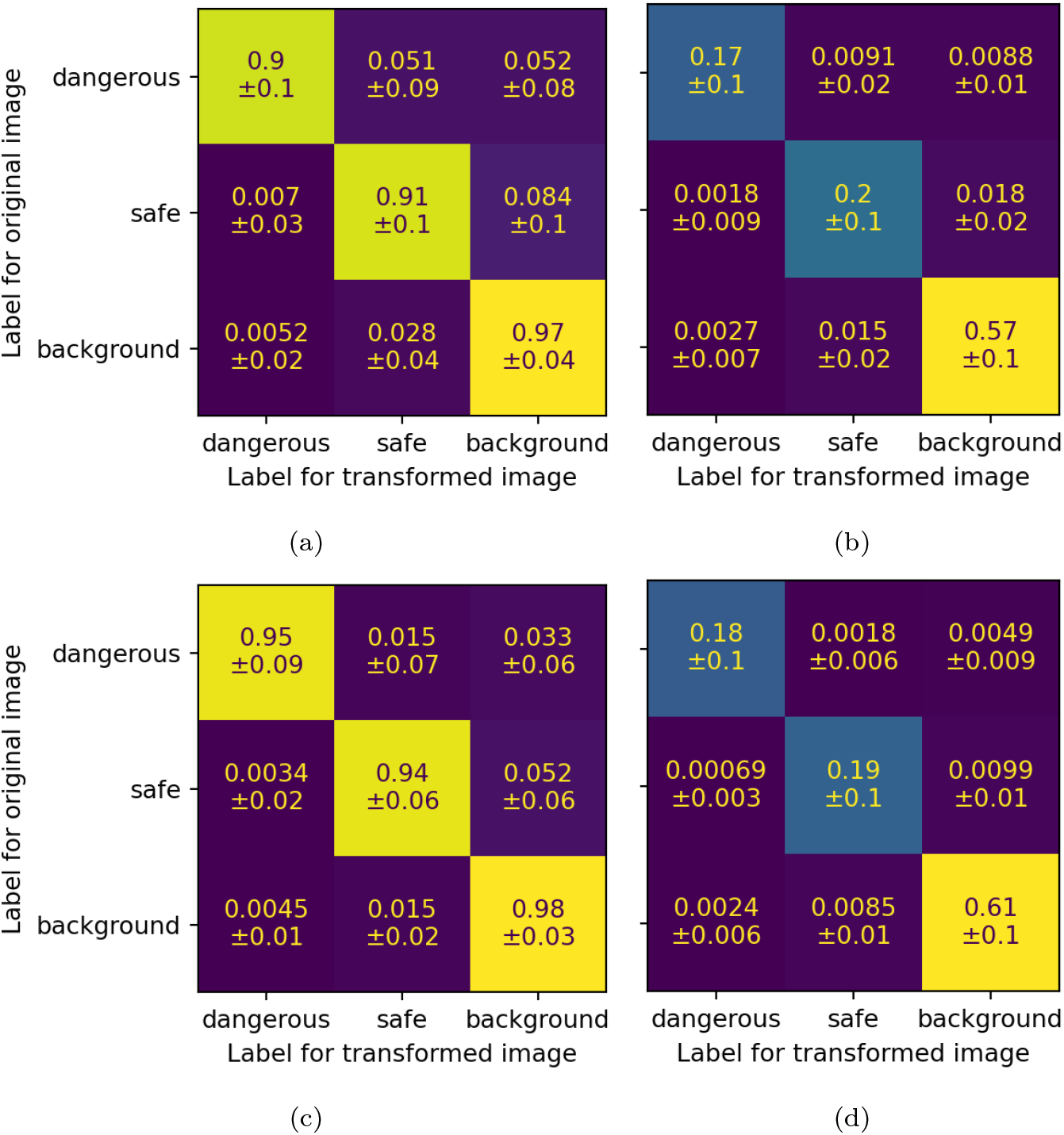
Confusion matrices for lighting bias. a.) Row-normalized, without tool pasting or lighting augmentations, b.) Fully-normalized, without tool pasting or lighting augmentations, c.) Row-normalized, with lighting augmentations, d.) Fully-normalized, with lighting augmentations

### 4.6. Lighting bias mitigations

Row-normalized and fully-normalized confusion matrices for the best model trained with random lighting augmentations are presented in Figure 8c and Figure 8d. They show that only 1.5% (was 5% without augmentations) of dangerous zone pixels can now be made to be predicted as safe zone, and 3.3% (was 5%) – as background. A model that uses lighting augmentation also results in much more consistent sizes of predicted zones (dangerous zone shrinks by 2%, safe zone shrinks by around 0.1% and background grows by less than 0.7%). Random lighting augmentation makes the model more robust to lighting changes, and helps prevent dangerous pixel misclassifications.

### 4.7 Segmentation performance

While we have shown that models with lighting and tool augmentations are more consistent, it is also important to validate that their performance on the core task has not degraded as a result. Figure 9 shows row-normalized confusion matrices for segmentation quality for the baseline model and model with both two iterations of tool pasting augmentation and lighting augmentation. These confusion matrices show that the two models perform roughly on the same level in terms of segmentation quality, with the model with the augmentations performing slightly better, demonstrating that the proposed tool pasting and lighting augmentations do not affect segmentation quality on the dataset affected by both tool and location biases. One might wonder why the segmentation performance is almost unchanged given that the models trained with augmentations are supposed to generalize better by being more robust to spurious correlations. The reason is that such models are only supposed to generalize better to datasets not affected by these correlations, which, in this case, means real-life surgeries performed by less experienced surgeons, who may have difficulties with equipment positioning. Since the entirety of our dataset, including training, validation and testing splits, has such shortcuts, we expected the segmentation performance to remain roughly on the same level.

**Figure 9:**
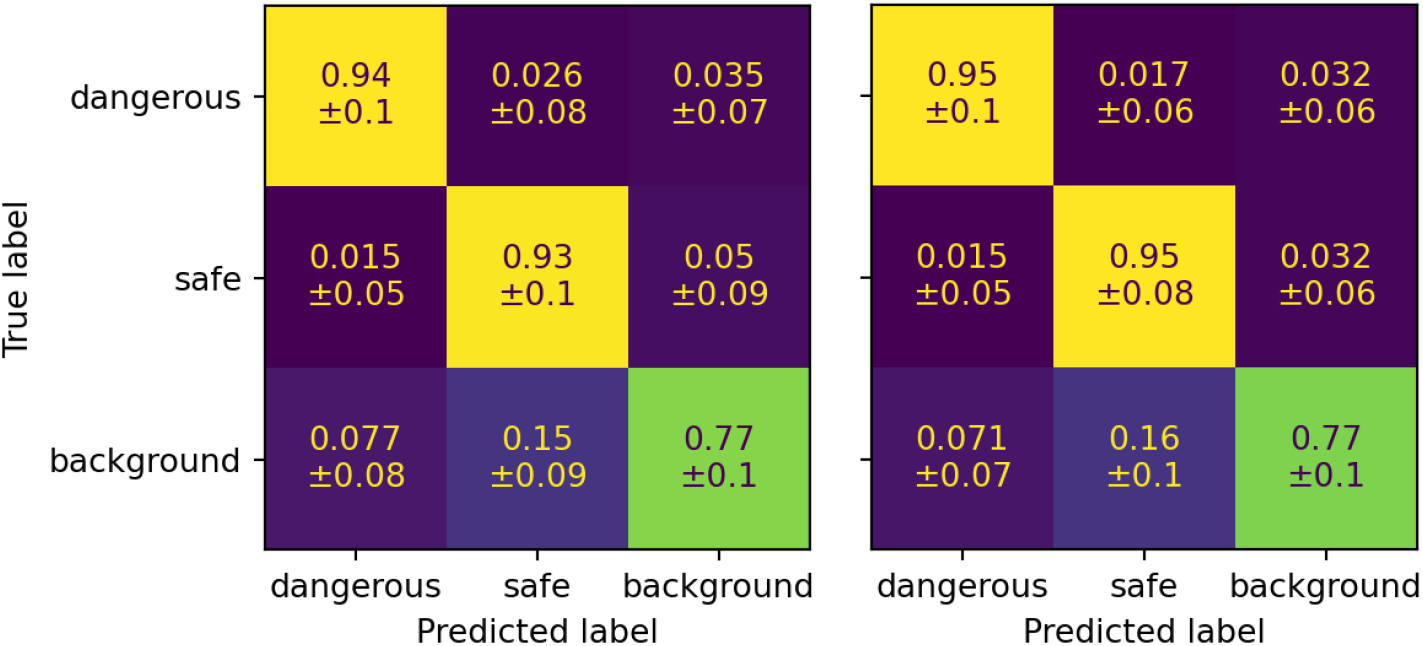
Row-normalized confusion matrices for segmentation quality. a.) Baseline model, b.) Model with both two iterations of tool pasting and lighting augmentations

## 5. Conclusions

In this work we considered the problem of surgical tool positions and lighting direction serving as confounders in the training data for surgical computer vision models for laparoscopic cholecystectomy. Data confounders in general and those related to surgical equipment configuration specifically pose a significant technical challenge, because they can’t be evaluated and mitigated by the widely-used practices in machine learning, such as training-validation-testing data splits. This is because all the data samples are affected by these shortcuts, so even evaluation on a held-out test set, even if it comes from a different institution, does not reveal model biases acquired during training.

The clinical significance of our work is particularly pronounced when considering the human factors inherent to surgical practice. Surgeons, especially those in training or practicing in resource-limited settings, may not consistently position equipment in the optimal manner that is typical of expert surgeons in high-volume centers. An AI system that has learned to rely on these expert-specific equipment placements as shortcuts for identifying critical anatomy could provide dangerously misleading guidance when used by less experienced operators. Our mitigation strategies specifically address this safety concern by decoupling anatomical recognition from equipment positioning, creating models that provide reliable guidance regardless of the surgeon’s technique or expertise level.

While we only considered the effect that these confounders have on models for identification of safe and dangerous zones of dissection in the context of LC, we believe that both the problem we tackled and the proposed solutions should generalize both to other tasks for LC datasets, such as identification of liver, gallbladder, hepatocystic triangle, or assessment whether the established field of view and retraction are adequate, and to other surgical computer vision problems. For both tool and location biases we proposed augmentation-based mitigation strategies that help train much more robust models. Being data-driven and model-agnostic, these strategies should generalize beyond the SegFormer architecture that we used for our experiments. Analysis of segmentation quality of models trained with these augmentations shows that their performance remains on the same level when evaluated on a biased dataset.

From a data perspective, our work highlights a fundamental challenge in medical AI development: the pervasive nature of shortcuts in surgical video datasets that are not addressed by simple train-validation-test splits. Even with carefully partitioned data, models can learn spurious correlations that persist across all splits because they reflect inherent patterns in how procedures are performed. Our evaluation methodology reveals that standard segmentation metrics like Dice score can obscure these critical reliability issues, as models may achieve high performance while remaining vulnerable to equipment-induced failures. This underscores the need for more robust evaluation frameworks in medical computer vision that specifically test for consistency across variable conditions, not just accuracy on static test images. The data augmentation strategies we propose represent a practical approach to breaking these correlations during training, but they also highlight the broader need for more diverse surgical datasets that explicitly capture variability in equipment positioning and other technical factors that could otherwise become hidden confounders.

## Data Availability

All data used in this research is either already available and cited appropriately, or available upon request.

## 6. Acknowledgments and conflicts of interest

This study was funded by the University Health Network Foundation and an NSERC Discovery grant to M.B. M.B. is a CIFAR CCAI chair. Other authors declare that they have no known competing financial interests or personal relationships that could have appeared to influence the work reported in this paper.

## 7. Data availability

- Dataset previously referred to as “Dataset 1” in [18] is available upon request through the Global Surgical Artificial Intelligence Collaborative. For access, please.
- CholecTrack20[30] dataset is available for download from Synapse, as described [here](https://github.com/CAMMA-public/cholectrack20#download)
- Endoscapes[31] dataset is available for download as described [here](https://github.com/CAMMA-public/Endoscapes#overview)

## 8. Declaration of generative AI and AI-assisted technologies use in the manuscript preparation process

During the preparation of this work the authors used a GLM 4.6 model as provided by Venice AI service for editorial assistance. After using this service, the authors reviewed and edited the manuscript and take full responsibility for the content of the published article.

## 9. Author roles

- **Conceptualization:** Protserov, Madani, Brudno
- **Data curation:** Protserov, Repalo, Hunter
- **Formal analysis:** Protserov, Hunter, Brudno
- **Funding acquisition:** Madani, Brudno
- **Methodology:** Protserov, Repalo, Mashouri, Hunter, Brudno
- **Project administration:** Masino
- **Software:** Protserov, Repalo, Mashouri, Hunter
- **Supervision:** Madani, Brudno
- **Writing – original draft:** Protserov, Brudno
- **Writing – review and editing:** all authors

